# Surgical management of hepatic cystic echinococcosis – a 20-year case series and outcome analysis

**DOI:** 10.1101/2025.10.20.25338349

**Authors:** I Barreto, A Deibel, J Fehr, B Müllhaupt, A E. Kremer, H Petrowsky, J Oberholzer, JP Jonas

**Affiliations:** Department of Visceral and Transplant Surgery, University Hospital Zurich, Zurich, Switzerland; Department of Gastroenterology and Hepatology, University Hospital Zurich, University of Zurich, Zurich, Switzerland

## Abstract

**Background:** Cystic echinococcosis (CE) is a parasitic zoonosis, caused by the larval stage of *Echinococcus granulosus*, typically forming cysts in the liver. If left untreated, CE can be life-threatening. Albendazole remains the standard medical therapy, but surgical intervention is indicated in selected cases.

**Methodology/Principal Findings:** We conducted a retrospective, multicenter outcome analysis in Switzerland of surgically treated hepatic cystic echinococcosis from 2004 to 2024. Thirty-one patients with CE stages I-III who underwent pericystectomy, endocystectomy, or minimally invasive procedures were included. The primary objective was to evaluate the safety and efficacy of surgical treatment. Data on patient demographics, disease characteristics, and perioperative morbidity and mortality, as well as recurrence rates stratified by surgical technique were analyzed.

All patients received adjuvant albendazole therapy for a median duration of 4 months (3–204 months). Across 32 surgical interventions, most patients (81.3%; n=26) had one or two cysts (1– 8), with a median cyst size of 8 cm (3–19 cm). The median follow-up was 78 months (12–230 months). The overall recurrence rate was 6.3% (n=2); however, there were no recurrences observed after pericystectomy (n= 0) or endocystectomy (n=0). The 90-day postoperative mortality rate was 0%.

**Conclusions:** In combination with established pharmacological therapy, surgery remains a safe and effective treatment option for CE, associated with low recurrence and negligible perioperative mortality. Surgical therapy should be considered a valuable component of CE management, especially in patients who are not optimal candidates for antihelmintic therapy alone.

**Author Summary:** Cystic echinococcosis is a parasitic disease that causes cystic lesions, most commonly in the liver. Although antiparasitic medication plays an important role, surgery remains the only curative option in selected cases. A key surgical question concerns the extent of resection— whether to remove only the cyst contents and inner layers or to perform a complete excision of the cyst, including its pericystic wall. This decision has important implications for radicality, preservation of healthy tissue, and the risk of recurrence.

We conducted a multicenter, retrospective analysis covering 20 years of surgical treatment for cystic echinococcosis at a Swiss tertiary referral center and partner hospitals. Among 31 patients undergoing 32 procedures, pericystectomy was the most common technique. This method achieved complete removal of the cyst and was associated with no recurrences during long-term follow-up, whereas more conservative approaches showed isolated relapses. Overall morbidity was low, and there was no procedure-related mortality.

These findings support pericystectomy, combined with perioperative anthelmintic therapy, as an effective and safe standard of care for selected patients requiring operative treatment.

## Introduction

Cystic echinococcosis (CE) is a zoonotic disease caused by the larval stage of the dog tapeworm *Echinococcus granulosus sensu lato*, which comprises five species and at least ten genotypes. Although these parasites occur worldwide, the disease is primarily endemic in developing and emerging countries. Humans become infected through fecal–oral transmission and represent accidental intermediate hosts (1). The liver is the main organ affected, although cysts may develop in nearly any organ, including the lungs, spleen, kidneys, and brain (2,3). Up to 80% of patients present with a solitary cyst involving a single organ, most commonly the liver (80%) or lungs (20%).

In contrast to *E. multilocularis*, the larval tissue of *E. granulosus* grows by displacement rather than infiltration (2). Consequently, CE is more amenable to surgical or minimally invasive treatment, and its prognosis is significantly better than that of alveolar echinococcosis (AE) (4,5). The most widely used minimally invasive technique is PAIR (puncture, aspiration, injection, and reaspiration), which aims to destroy the germinal layer. PAIR is suitable for CE1 and CE3a cysts but carries risks such as anaphylaxis and recurrence (8,17). Alternatively, the modified catheterization technique (MoCT) can be used to evacuate the entire endocyst (4). This method is reserved for large cysts or those containing daughter vesicles (CE2, CE3b) and is associated with risks including secondary bacterial infection, abscess formation, or biliary fistula. Both procedures are primarily employed in endemic regions such as Turkey (9), while expertise is often limited in non-endemic countries like Switzerland.

Surgical indications traditionally include cyst-related complications such as rupture, biliary fistula, compression of vital structures, and secondary infection or hemorrhage (16). Surgery is also indicated for cysts containing multiple daughter vesicles that are unsuitable for percutaneous treatment (e.g., WHO stages CE2 and CE3b) (4,9). Additional indications include cysts larger than 10 cm and superficial cysts at risk of rupture due to trauma (4).

Adjunctive antiparasitic therapy is mandatory to reduce the risk of secondary echinococcosis from intra-abdominal seeding in the event of cyst fluid spillage (6).

Curative treatment of CE requires complete excision of the cyst, including all structural components—germinal, laminated, and adventitial layers. Incomplete removal, as in endocystectomy, partial cystectomy, or percutaneous approaches, carries a significant risk of recurrence. However, the optimal extent of surgical resection remains debated. It is still unclear whether formal hepatectomy is necessary in all cases or whether total cyst excision (pericystectomy) achieves comparable outcomes. Hepatectomy may reduce recurrence risk but involves resection of functionally intact liver parenchyma, increasing perioperative morbidity (10–13). In contrast, pericystectomy allows radical cyst removal, including the adventitial layer, while preserving healthy liver tissue, potentially balancing efficacy and safety.

Recurrence rates after resection range from 2% to 25% (7–9,15), depending on cyst location, size, and surgical expertise.

This multicenter analysis aims to evaluate surgical management of hepatic cystic echinococcosis with respect to safety and recurrence outcomes.

## Methods

### Study Design

A retrospective cohort study was conducted on patients with hepatic cystic echinococcosis who underwent surgical resection or minimally invasive procedures at the University Hospital Zurich and its partner cantonal hospitals between 2004 and 2024. Thirty-one patients were included, all of whom provided written informed consent. The study protocol was approved by the Ethics Committee for Human Research in Zurich (BASEC Number: 2024-01732). Among the 31 patients, one underwent reoperation for recurrence following open cyst aspiration and fenestration, resulting in a total of 32 interventions.

The primary aim of this study was to analyze the morbidity, mortality, and recurrence risk associated with the surgical treatment of hepatic cystic echinococcosis. The outcome measures included the Dindo-Clavien classification of complications, the Comprehensive Complication Index (CCI) at discharge, length of hospital stay, 30-day mortality, overall survival, and recurrence rate (11, 12, 13). The secondary objectives were to assess factors influencing recurrence and complications, such as patient comorbidities, surgical techniques, cyst size, cyst number, and biliary involvement. Inclusion criteria consisted of radiologically confirmed hepatic E. granulosus infection with WHO stage classification CE I-III. The indication for surgery was made through a multidisciplinary approach. The diagnosis of cystic echinococcosis was established using serologic testing via enzyme-linked immunosorbent assay (ELISA) and imaging studies such as ultrasonography (US) and computed tomography (CT). CT imaging evaluated vascular involvement, while magnetic resonance imaging (MRI) was employed during the preoperative evaluation of biliary tract involvement. Radiological assessments documented cyst number, size, and WHO classification. Data extracted from patient records included patient characteristics (sex, age, cardiac, pulmonary, and renal comorbidities, bleeding disorder, nicotine use, Eastern Cooperative Oncology Group (ECOG) performance status, and American Society of Anesthesiologists (ASA) classification), disease characteristics (cyst size, cyst number, biliary involvement, vascular compression, anthelmintic medication, and its duration), as well as surgical data (technique, duration, blood loss, pringle maneuver, water-jet dissection, irrigation with saline or glucose solution, cyst opening, CCI at discharge, and 90-day mortality). Comorbidities were defined as diagnoses related to organ dysfunction, regardless of whether pharmacological treatment was given. Cardiac comorbidities included arterial hypertension, coronary artery disease, and dysrhythmias. Pulmonary comorbidities included chronic obstructive pulmonary disease, restrictive lung diseases, and previous lung resections that led to reduced respiratory capacity. Renal comorbidities were identified as chronic kidney disease with a reduced glomerular filtration rate. Recurrence was defined as the appearance of new active cysts after treatment, including regrowth of live cysts at the site of a previously treated cyst or the development of new distant disease due to cyst spillage.

Postoperative management and follow-up were standardized and conducted routinely. At our center, Albendazole is generally administered starting at least one week prior to surgery and continued for a minimum of three months postoperatively. The protocol includes serologic testing (ELISA) and abdominal ultrasonography at six months, one year, and annually thereafter in the absence of recurrence. A minimum follow-up of 10 years is performed at our center unless the patient declines. Any suspicion of recurrence identified on ultrasonography is confirmed with CT imaging and first managed pharmacologically.

### Surgery

Two techniques to surgically address the hepatic CE cyst were utilized. Pericystectomy is usually performed with the assistance of a parenchyma dissection device such as CUSA or Water-Jet. In this technique, surgeons operate along the outer layer of the cyst and dissect the hepatic parenchyma away from it using these tools. This can achieve complete clearance of the cyst from pedicles without the need for primary liver resection, resulting in maximal parenchyma sparing. Pericystectomy is, by definition, associated with technically R0 resection margins if performed correctly, as the dissected parenchyma provides a safety margin of a few millimeters. On the other hand, endocystectomy involves surgically opening the cyst with precautionary measures placed around the liver (e.g., gauzes saturated with hypertonic saline) to prevent content contamination. After the cyst is opened, the contents are carefully aspirated, and the cavity is irrigated with saline. This may be followed by enlarging the opening and removing the germline layer to prevent recurrence and eliminate any infectious content. This surgery is, by definition, not an R0 procedure because the outermost layer of the CE cyst still remains in situ and is therefore associated with a certain risk of recurrence.

### Statistical Analysis

All data were recorded using IBM SPSS software, version 16 for Mac (SPSS Inc.). A descriptive analysis of patient characteristics (sex, age, cardiac, pulmonary, and renal comorbidities, bleeding disorders, nicotine use, ECOG performance status, and ASA classification), disease characteristics (cyst size, cyst number, biliary involvement, vascular compression, anthelmintic medication, and its duration), and surgical data (technique, duration, blood loss, pringle maneuver, water-jet dissection, irrigation with saline or glucose solution, cyst opening, CCI at discharge, and 90-day mortality) was performed.

For the primary outcomes, postoperative complications were analyzed using the Dindo-Clavien classification and the Comprehensive Complication Index (CCI), reported as frequencies and medians, respectively. Recurrence rates were calculated as proportions and stratified by surgical technique. Overall survival was analyzed using the Kaplan-Meier survival curve.

All analyses were descriptive and exploratory in nature, reflecting the limited sample size and number of events.

## Results

### Patient Demographics

The majority of patients, 58.1% (n=18), were male, with a median age of 45 years (28 - 80). A total of 90.4% (n=28) of patients came from echinococcosis-endemic areas (Kosovo, Macedonia, Tunisia, Turkey, Italy, Bulgaria, Montenegro, Syria, China), and 16.1% (n=5) had previously undergone parenchyma-sparing surgery for CE in their respective countries before arriving in Switzerland. Cardiac comorbidities were the most frequently observed (19.4%), followed by renal comorbidities (9.7%) and pulmonary comorbidities (3.2%). Fifteen patients (48.4%) were active smokers at the time of surgery. The ECOG performance status was <2 in 96.8% of patients, and the ASA score was predominantly 2 (87.1%). The demographic characteristics of all patients are summarized in Table 1.

**Table 1:** Demographic characteristics of the patient population.

|  | **Patients with**<br>**hepatic cystic**<br>**echinococcosis**<br>**N=31** |
| --- | --- |
| **Sex** |  |
| Male | 18 (58.1 %) |
| Female | 13 (41.9 %) |
| **Age (median)** | 45 (28 - 80) |
| **Country of Origin** |  |
| Kosovo | 7 (22.6 %) |
| Macedonia | 6 (19.4 %) |
| Tunisia | 2 (6.5 %) |
| Turkey | 7 (22.6 %) |
| Italy | 1 (3.2 %) |
| Bulgaria | 1 (3.2 %) |
| Montenegro | 1 (3.2 %) |
| Switzerland | 3 (9.7 %) |
| Syria | 3 (9.7 %) |
| **Cardiac Co-morbidities** | 6 (19.4 %) |
| **Pulmonary Co-morbidities** | 1 (3.2 %) |
| **Renal Co-morbidities** | 3 (9.7 %) |
| **Bleeding disorder** | 2 (6.5 %) |
| **Smoking status** |  |
| Current smokers | 15 (48.4 %) |
| Past smokers | 1 (3.2 %) |
| Non-smokers | 15 (48.4 %) |
| **ECOG** |  |
| 0 | 25 (80.7 %) |
| 1 | 5 (16.1 %) |
| 2 | 1 (3.2 %) |
| 3 | 0 (0.0%) |
| 4 | 0 (0.0 %) |
| 5 | 0 (0.0 %) |
| **ASA** |  |
| 1 | 3 (9.7 %) |
| 2 | 27 (87.1 %) |
| 3 | 2 (6.5 %) |

### Disease characteristics

Most patients had one (65.6%, n=21) or two (15.6%, n=5) cysts, and the median cyst size was 8 cm (3–19 cm). All patients received pharmacological treatment perioperatively, either with albendazole (87.5%) or mebendazole (12.5%). The median duration of perioperative pharmacological treatment was 3 months (2-94). Biliary tract compression was observed in 6.3% (n=2) of cases, and vascular compression (of the right hepatic vein, middle hepatic vein, and/or inferior vena cava) was noted in 18.8% (n=6) of cases. The data regarding disease characteristics are detailed in Table 2.

**Table 2:**
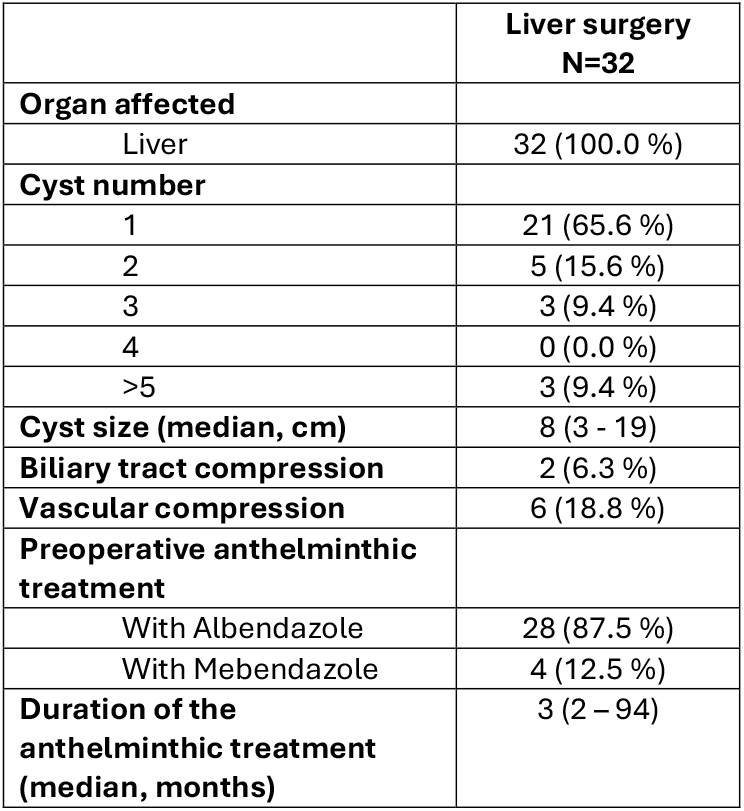
Disease characteristics of the patient population.

### Surgery

Open pericystectomy was the most commonly performed procedure overall, accounting for 84.4% (n=27), followed by open endocystectomy at 9.4% (n=3). Among the 27 pericystectomies, 16 procedures were limited resections, 2 were bisegmentectomies, 6 were right hepatectomies, and 3 were left hepatectomies (see Table 3). For the resective procedures (n=27), an R0 resection rate of 96.3% was observed (see Table 4). In the only R2 case, no disease remained in the liver; however, the disease was present in another anatomical region, deemed inoperable, specifically in the cervical spine. In this case, a 9.4 cm cyst in the liver necessitated a bisegmentectomy of segments IVa and VIII due to its size and location. The median duration of surgery was 307.5 minutes (142-600 minutes), and the median blood loss was 290 ml (95-2000 ml). The Pringle maneuver was performed in 43.8% (n=14) of cases, with a documented total duration ranging between 15 and 60 minutes. The dissection was conducted using a water-jet technique in 46.9% of cases (n=15). A controlled cyst opening rate of 40.6% was observed. Surgeons utilized a hyperosmolar saline solution of 40% in 34.4% (n=11) of all cases or a 40% glucose solution in 6.3% (n=2) of cases. All surgical parameters are presented in Table 3.

**Table 3:**
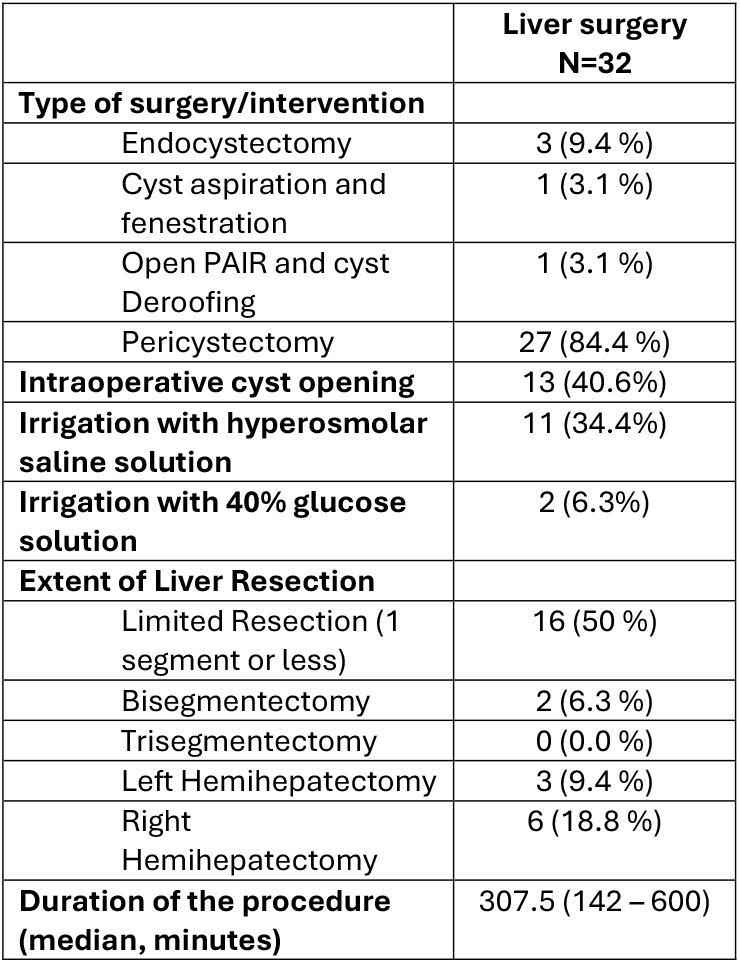
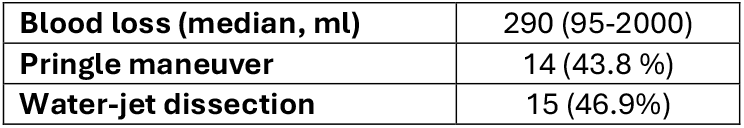
Surgical parameters.

**Table 4:** Resection Status.

|  |  |
| --- | --- |
|  | Hepatic resective procedures (n= 27) |
| **Resection Status** |  |
| R0 | 26 (96.3 %) |
| R2 | 1 (3.7 %) |

### Postoperative course

Twelve patients (37.5%) were transferred to the intensive care unit postoperatively. The median length of hospital stay was 8 days (5 - 23). Overall complications occurred in 40.6% (n=13) of cases; however, only 5.5% (n=1) of all complications (n=18) were severe (Clavien-Dindo IIIb). The median Comprehensive Complication Index (CCI) was 13.3 (0 – 47.2) at discharge. Reported complications included pneumothorax and pleural effusion requiring chest tube insertion, pneumonia, urinary tract infection, bleeding necessitating transfusion, perihepatic fluid collection requiring percutaneous drainage, exudative pancreatitis, bacteriemia, pulmonary edema, atrial flutter, wound infection, and paralytic ileus. Iatrogenic pneumothorax, pneumonia, bleeding and paralytic ileus were the most common complications. No relaparotomy was necessary in the acute postoperative period. Accidental intraoperative cyst perforation occurred in one case, as did intraoperative diaphragmatic perforation in another case. See Table 5 for a detailed description of the reported postoperative complications. The 90-day mortality rate was 0%, and the only fatality in the cohort occurred four years later due to complications from a heart transplant. Table 6 provides further information regarding primary and secondary outcomes.

**Table 5:** Reported postoperative complications.

| **Postoperative Complication** | **Number of events reported** |
| --- | --- |
| Iatrogenic Pneumothorax | 2 |
| Pneumonia | 2 |
| Pleural effusion | 1 |
| Urinary Tract Infection | 1 |
| Bleeding | 2 |
| Perihepatic fluid collection | 1 |
| Exudative Pancreatitis | 1 |
| Bacteremia | 1 |
| Pulmonary Edema | 1 |
| Atrial Flutter | 1 |
| Wound infection | 1 |
| Paralytic ileus | 2 |
| Accidental intraoperative cyst perforation | 1 |
| Intraoperative diaphragmatic perforation | 1 |

**Table 6:**
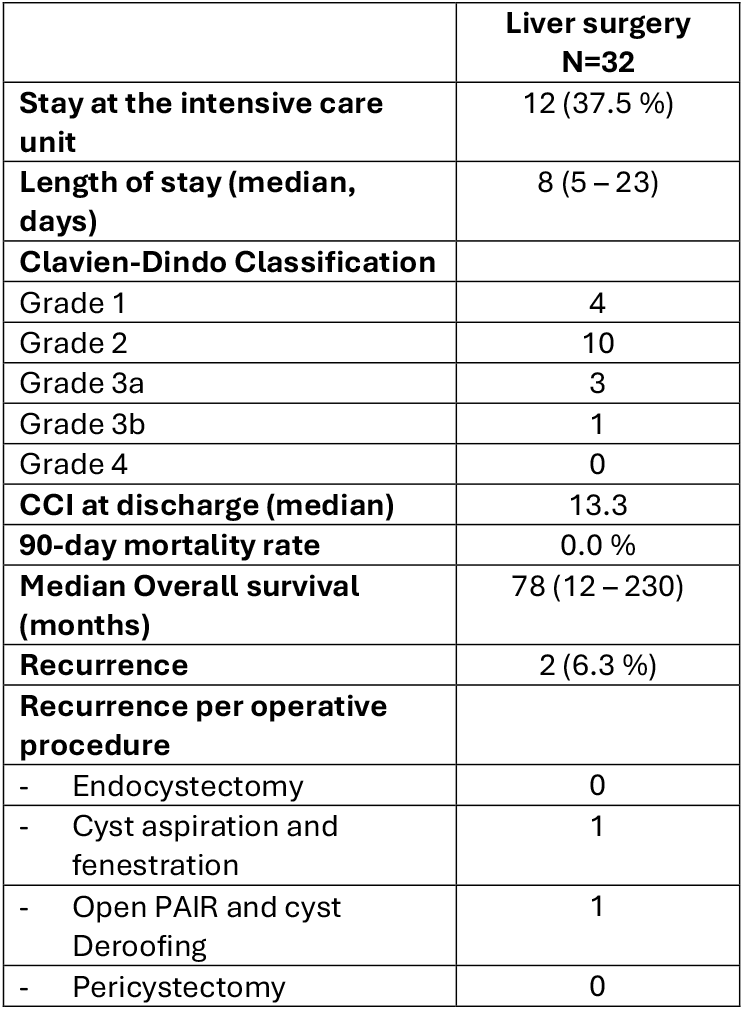
Primary and secondary outcomes.

|  | **Liver surgery**<br>**N=32** |
| --- | --- |
| **Stay at the intensive care unit** | 12 (37.5 %) |
| **Length of stay (median, days)** | 8 (5 – 23) |
| **Clavien-Dindo Classification** |  |
| Grade 1 | 4 |
| Grade 2 | 10 |
| Grade 3a | 3 |
| Grade 3b | 1 |
| Grade 4 | 0 |
| **CCI at discharge (median)** | 13.3 |
| **90-day mortality rate** | 0.0 % |
| **Median Overall survival (months)** | 78 (12 – 230) |
| **Recurrence** | 2 (6.3 %) |
| **Recurrence per operative procedure** |  |
| - Endocystectomy | 0 |
| - Cyst aspiration and fenestration | 1 |
| - Open PAIR and cyst Deroofing | 1 |
| - Pericystectomy | 0 |

### Follow-up

The median follow-up time was 78 months (12–230 months). During this period, two cases of recurrence (6.3%) were identified, necessitating repeated resection in one of the cases. The patient, who had gone through open cyst aspiration and fenestration, developed a new retention cyst in liver segments VII/VIII, which compressed the vena cava. Consequently, a relaparotomy with right hemihepatectomy was needed seven months later. In the other case, a hepatic recurrence was noted three months after open PAIR and cyst deroofing, which was treated pharmacologically with albendazole. Overall survival and recurrence-free survival by surgical technique are shown in Figure 1 and Figure 2, respectively.

**Figure 1.**
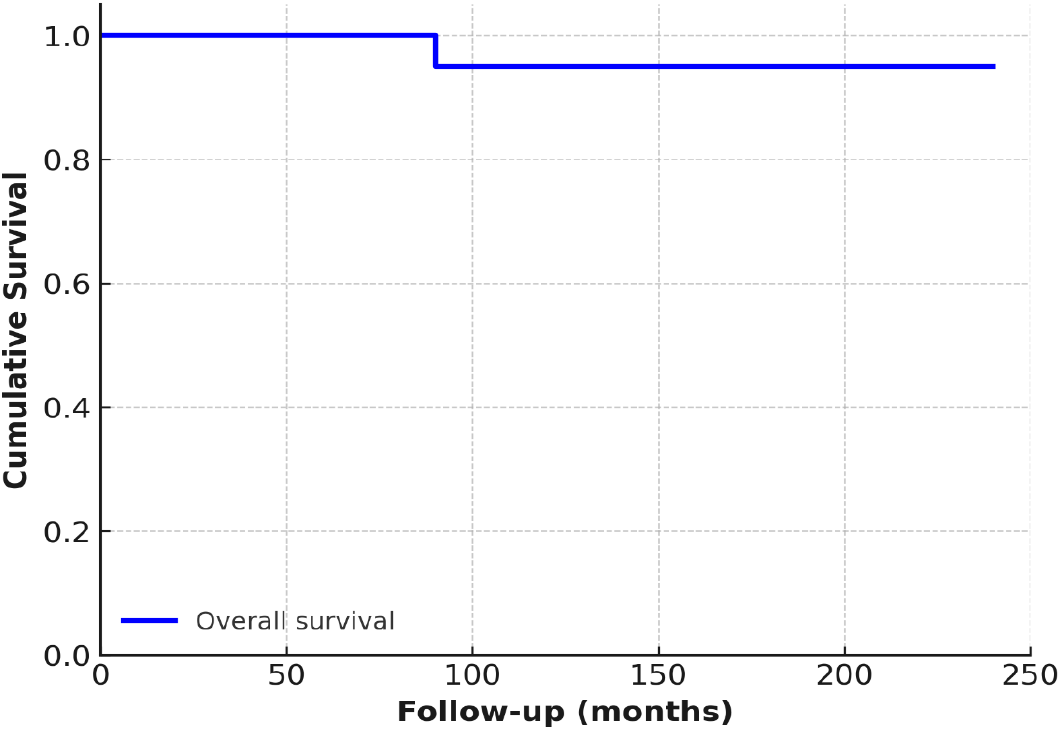
Kaplan-Meier overall survival curve

**Figure 2.**
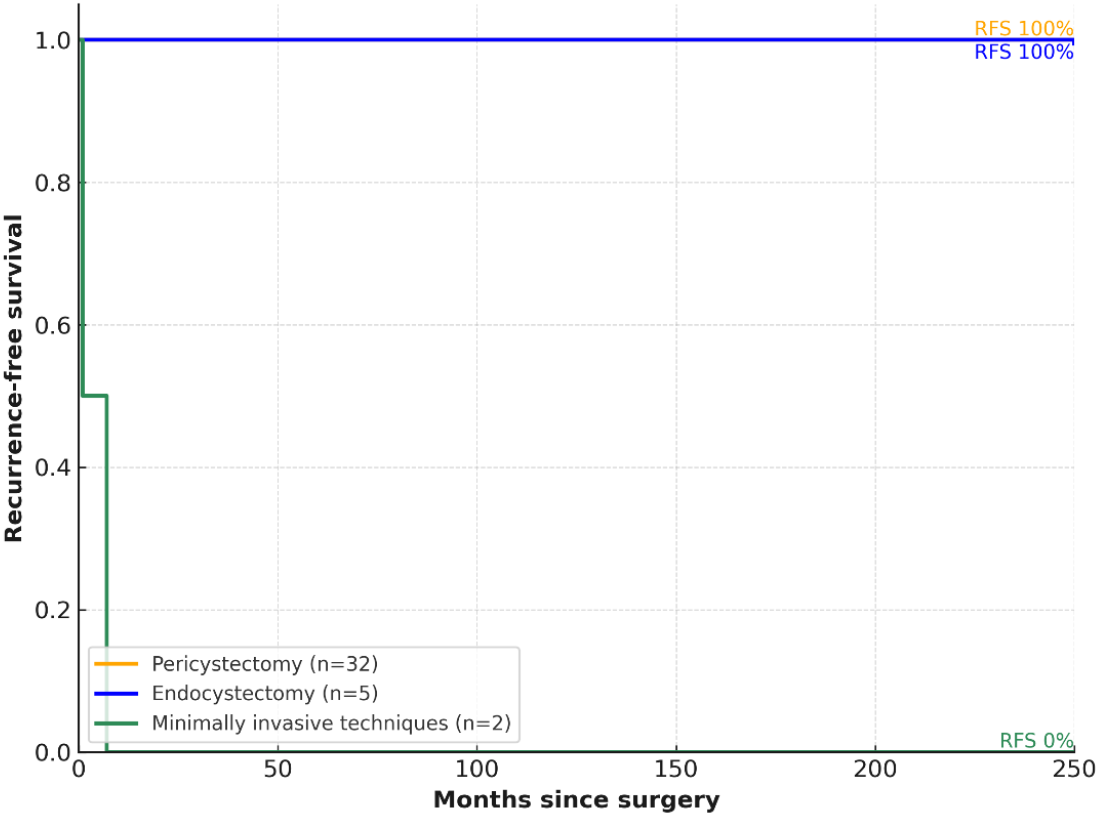
Recurrence-free survival by surgical technique

## Discussion

This retrospective cohort study examines 20 years of surgical management for hepatic cystic echinococcosis at a Swiss tertiary referral center and partner cantonal hospitals, offering significant insights into long-term outcomes in a non-endemic environment. Among the 31 patients who underwent surgical treatment, most received pericystectomy or endocystectomy. The recurrence rate was 6.3%, occurring solely in the minimally invasive treatment subgroup, while no recurrences were observed in the endocystectomy and pericystectomy groups. Postoperative complication rates were low, with no 90-day mortality reported. These findings confirm that surgical treatment, particularly pericystectomy, remains a safe and effective option when indicated, even in regions with limited endemic exposure (18).

For CE, the primary surgical techniques are pericystectomy and endocystectomy, each presenting unique benefits and drawbacks. Pericystectomy involves the complete excision of the hydatid cyst, including both the parasitic components and the host-derived adventitial layer. It is classified as a radical procedure and has been consistently associated with lower recurrence rates and better long-term outcomes. Cirenei et al. noted a recurrence rate of just 0.9% following radical procedures like pericystectomy or liver resection, as opposed to 11.2% for patients receiving conservative surgery such as endocystectomy (20). Rinaldi et al. further emphasized that pericystectomy significantly reduces the risk of postoperative complications related to residual cavity infections or biliary fistulas, due to the thorough removal of infected and inflamed tissues (21). Govindasamy et al. also observed that recurrence is minimized when the entire pericyst is excised, particularly by preventing exogenous vesiculation and secondary cyst formation (22). However, pericystectomy remains technically demanding and carries increased intraoperative risk, especially when cysts are attached to major vascular or biliary structures (19).

Endocystectomy is a parenchyma-sparing technique that removes the internal parasitic germinal layer while retaining the fibrous host capsule, which may contain viable scolices and contribute to recurrence. A recent systematic review encompassing 54 studies (n = 4.058 patients) indicated a recurrence rate of 4.8% and a mortality rate of 1.2% for endocystectomy, with a higher likelihood of recurrence if cavity management is not optimal (23). Although these outcomes may be considered acceptable, particularly in resource-limited settings or anatomically complex situations, they are significantly inferior to those achieved through pericystectomy regarding recurrence prevention. The elevated recurrence risk of endocystectomy has been associated with exogenous vesiculation and the challenges of ensuring complete removal of germinal debris from the residual cavity (23).

A valuable adjunct to pericystectomy that enhances safety and precision is the use of water-jet dissection. This technique facilitates selective parenchymal dissection while protecting biliary and vascular structures, which significantly reduces the risk of bleeding and bile leaks. In our institution’s experience, water-jet dissection enables *en bloc* removal of the cyst with minimal tissue damage, especially in lesions that are centrally or perivascularly located. Although prospective comparative studies are limited, emerging evidence highlights the utility of water-jet dissection in hepatobiliary surgery for CE, particularly in enhancing the thoroughness of pericystectomy while maintaining surgical safety.

In conclusion, optimal CE treatment should be multimodal and tailored to individual patient characteristics. Radical pericystectomy should be considered the standard treatment for surgically suitable patients, owing to its superior long-term outcomes and significantly lower recurrence rates, according to the literature. Endocystectomy may still be necessary for high-risk patients or when radical resection is unfeasible due to anatomical constraints. Adjunctive techniques like water-jet dissection enhance the safety and feasibility of pericystectomy and should be utilized in complex cases. Antiparasitic therapy with albendazole remains essential before and after surgery. Ultimately, a multidisciplinary and staged approach—including surgery, medical therapy, and, where appropriate, endoscopic or percutaneous methods—provides the best opportunity for durable disease control and minimizing recurrence in CE.

## Conclusion

Surgical therapy continues to be performed globally for specific indications of CE cysts. This case-series highlights that surgery remains a cornerstone of multimodal CE therapy, with negligible procedure-related mortality. Although complications are possible, serious complications rarely occur. Unlike PAIR or MoCAT, surgery allows complete cyst removal, including the germinal layer, in a single intervention and is associated with negligible recurrence rates. In our cohort, pericystectomy and endocystectomy demonstrated a 0% recurrence rate with a very low procedure-related risk.

## Data Availability

All relevant data are within the manuscript and its Supporting Information files.

## Limitations

A key limitation of this retrospective study is selection bias, as non-surgically treated patients were not included. Variability in surgical techniques and postoperative care further limits comparability and conclusions regarding the most effective approach. Moreover, the relatively small cohort size precluded robust statistical analysis, and the findings should therefore be interpreted in a descriptive and exploratory context.

